# Choroidal Neovascularization as a Trigger for Central Serous Chorioretinopathy

**DOI:** 10.1101/2025.08.27.25334593

**Authors:** Aliénor Vienne-Jumeau, Elodie Bousquet, Jacques Bijon, Sarah Mrejen, Francine Behar-Cohen

## Abstract

**Purpose:** To investigate whether choroidal neovascularization (CNV) can act as a trigger for central serous chorioretinopathy (CSCR) in patients with pachychoroid features, by analyzing cases where fluorescein angiography (FA)-confirmed leakage originated directly within the CNV lesion.

**Methods:** We retrospectively reviewed patients with no prior history or signs of CSCR who presented with a first episode of CSCR and coexisting CNV. Inclusion required at least one FA-confirmed leakage point located within the neovascular complex. Multimodal imaging— including FA, spectral-domain optical coherence tomography (SD-OCT), and OCT angiography (OCTA)—was performed to detect CNV and evaluate its topographical relationship with leakage.

**Results:** Among 202 patients screened, four met inclusion criteria (two males, two females; age range 54–58 years). All presented with a unilateral first episode of CSCR. In each case, FA and OCTA demonstrated precise colocalization of the leakage point within the CNV lesion. CNV was predominantly retrofoveal and appeared mature in morphology. Three patients were diagnosed simultaneously with CSCR and a neovascular membrane, while one developed CSCR during follow-up of a previously identified PNV. Subretinal fluid fluctuations were observed in all cases and often appeared independent of anti-VEGF treatment, suggesting a mechanism not exclusively driven by VEGF-mediated CNV activity.

**Conclusions:** This case series suggests that, within the pachychoroid spectrum, CNV may not only complicate chronic or complex CSCR but may also act as a direct trigger of acute episodes. These findings underscore the importance of multimodal imaging, particularly FA and OCTA, for comprehensive assessment of CSCR.

## Introduction

Acute central serous chorioretinopathy (CSCR) is defined by the occurrence of serous retinal detachment associated with a leak on fluorescein angiography (FA), showing active dye passage through one or more areas of the retinal pigment epithelium (RPE) barrier break.(Spitznas and Huke, 1987), (Levine et al., 1989) Other characteristic clinical signs are visual impairment such as contrast loss, color vision impairment, micropsia, and focal blurring, without major loss of visual acuity.(Daruich et al., 2015), (Zhang et al., 2023) Spectral-Domain Optical Coherence Tomography (SD-OCT) examination confirms the diagnosis when characteristic choroidal signs are present, such as thickened choroid compared to fellow eye or age-matched controls and the presence of dilated veins that obscure the choriocapillaris (pachyvessels)(Daruich et al., 2017), (Orduña-Azcona et al., 2023) and are associated with the serous detachment. On SD-OCT, the leakage site(s) observed on FA often correspond to structural changes such as a pigment epithelial detachment (PED), focal RPE disruption, areas of outer segment ‘erosion’ or elongation(Daruich et al., 2015), (Li et al., 2022), and, in some cases, adjacent hyporeflective subretinal lucencies.(Bijon and Freund, 2024) In acute forms, FA typically reveals a focal site of RPE leakage. A high intensity of leakage is generally indicative of an episode that will resolve within a few months(Daruich et al., 2017). In contrast, diffuse RPE dysfunction without a clearly identified leakage site is more suggestive of chronic episodes and more complex disease forms(Parameswarappa et al., 2021). The exact mechanisms of “leakage,” which is transient in acute forms, are not fully understood. Histological analysis of adrenaline-induced CSCR-like disease in monkey showed that spots of fluorescein leakage coincided with focal RPE cell degeneration over area of choriocapillaris endothelial defects with fibrin deposits within the Bruch’s membrane, suggesting that coagulation cascade and macromolecular transports could be involved.(Yoshioka and Katsume, 1982a) Indocyanine green angiography (ICGA) in patients with acute CSCR confirmed choriocapillaris involvement with area of capillaries hypoperfusion and leakage at the site of RPE leak.(Iida et al., 1996; Piccolino et al., 1995) Several mechanisms have been hypothesized, including bulk fluid flow(Pryds et al., 2010) and reverse of retinal pigment epithelium pump(Fossataro et al., 2024), which could be promoted by mechanical stresses from the dilated and engorged choroid.

The development of choroidal neovascularization (CNV) is a sight threatening complication of CSCR, being responsible for a reduced visual acuity during a long-term follow-up.(Mrejen et al., 2019) Using multimodal imaging, CNV was detected in up to 40% of “chronic” or complex forms of CRSC.(Savastano et al., 2021) The detection of CNV can indeed be difficult, and it is the presence of a flat irregular PED (FIPED), partially hyperreflective, that should raise suspicion.(Bousquet et al., 2018a), (Azzolini et al., 2021), (Guo et al., 2021) Detection of choroidal neovascularization (CNV) is notably easier with OCT-Angiography (OCT-A), which enables visualization in approximately 35% of cases, compared to 25% with ICGA. This is because, unlike in age-related macular degeneration (AMD), type 1 CNV in these conditions does not frequently form hyperfluorescent plaques in the late phase of ICGA.(Zola et al., 2023) Although FIFED have been identified in around 40% of acute CSCR, the exact occurrence of CNV in these cases has been poorly documented. Within the pachychoroid spectrum, another entity has been recognized: pachychoroid neovasculopathy (PNV). These are patients who have no history of CSCR but present mature neovascularization secondary to pachychoroid and area of ICGA hyperpermeability, with or without associated epitheliopathy and no exudative detachment.(Pang and Freund, 2015) Differential diagnosis with AMD can be difficult, and it is the thickness of the choroid and the absence of drusen that will guide this diagnosis, the boundary with AMD currently being debated.(Miyake et al., 2015)

In this ever-expanding spectrum of pachychoroid disease, the definition of different forms of CSCR is an ongoing process.(Singh et al., 2018), (Sahoo et al., 2022) We report here the cases of patients presenting with a first episode of acute CRSC and an active FA leakage site located on the edge of a neovascular membrane. Based on these cases, we discuss the mechanisms, diagnostic methods, and implications for management.

## Methods

### Ethics Statement

The study was approved by the Ethics Committee of the French Society of Ophthalmology (00008855). It adhered to the tenets of the Declaration of Helsinki (1964). Oral informed consent was obtained from all patients prior to inclusion.

### Study Design

This retrospective observational study was conducted at the Department of Ophthalmology, Cochin Hospital (Paris), and the Department of Ophthalmology, Centre Hospitalier National d’Ophtalmologie des Quinze-Vingts (Paris). All patients aged ≥18 years who presented with either a new diagnosis or follow-up of central serous chorioretinopathy (CSCR) associated with choroidal neovascularization (CNV) between December 2024 and April 2025 were retrospectively reviewed for inclusion.

### Patients

Patients were eligible if diagnosed with acute CSCR associated with concurrent CNV. Inclusion criteria were: no prior history of CSCR, absence of clinical or imaging signs of complex CSCR, and presence of at least one leakage point located within the neovascular network. Multimodal imaging—including fluorescein angiography (FA), indocyanine green angiography (ICGA), spectral-domain optical coherence tomography (SD-OCT; Spectralis®, Heidelberg), and optical coherence tomography angiography (OCTA)—had to be performed within one month of CSCR diagnosis.

#### Data Collection

For each patient, the following data were collected: age at diagnosis, gender, CNV maturity, contralateral eye choroidal characteristics, and refractive error (spherical equivalent). CNV maturity was defined on OCTA as vascular morphology graded into immature (tangled or rosette-like cluster of indistinct capillary vessels) or mature (presence of a prominent, dilated core vessel within a more organized vascular network).(Uchida et al., 2019) The leaking point was defined as a focal area of hyperfluorescence with an increased in size over time on FA as previously described.(van Rijssen et al., 2019) CNV was defined on OCTA as abnormal flow within the outer retinal slab.(Kashani et al., 2017) When necessary, the outer retina and choriocapillaris slabs were manually adjusted to refine segmentation. CNV lesions were classified based on en face OCTA projections (ORCC slab) according to established vascular morphology criteria. Following identification of the leaking point and CNV, the OCTA was accurately realigned with the angiographic images by referencing the retinal vessels at the level of the superficial microvasculature, in order to precisely localize the site of leakage.

## Results

### Patients

Among 202 eligible patients with CSCR and CNV seen between December 2024 and April 2025, four patients were included (two males and two females; see Table 1). The age at CSCR onset ranged from 54 to 58 years. In all cases, only one eye was affected. The left eye was involved in two patients and the right eye in two. All patients were either emmetropic or mildly hyperopic, with absolute refractive errors <3 diopters.

### CSCR diagnosis and diagnosis in the Fellow Eye

All cases presented with a first episode of CSCR, which by definition manifests as serous detachment associated with an angiographic leaking point in the context of pachychoroid. No signs of chronicity or complexity were observed, such as outer retinal atrophy or widespread and/or multifocal RPE atrophy (≥2 disc diameters) (Figures 1A-F, 2A-D, 3A-D, 4A-D). The fellow eye of each patient showed distinct pachychoroid spectrum features. One patient had PNV, another had focal choroidal excavation (FCE), while the remaining two exhibited either pachychoroid pigment epitheliopathy (PPE) or uncomplicated pachychoroid. The patient with contralateral PNV manifest as a subtle flat irregular PED overlying a large and mature CNV and no sign or symptoms of an episode of subretinal fluid (Supplementary Figure 1).

**Figure 1.**
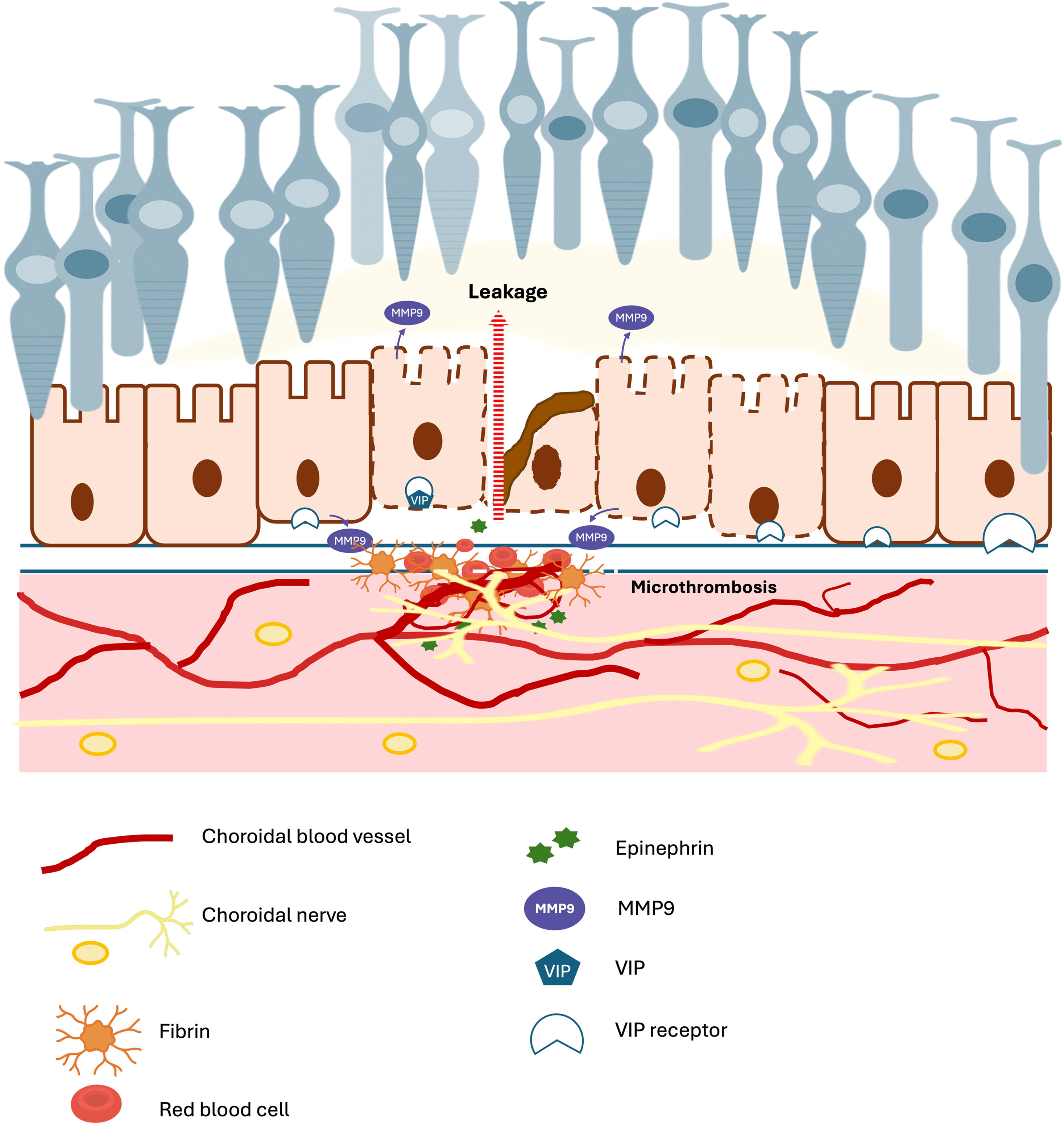
Multimodal follow-up imaging of the left eye in Case 1. Fluorescein angiography (FA) in the early, mid, and late phases (A–C) shows several hyperfluorescent pinpoint areas, including one with more intense leakage, while indocyanine green angiography (ICGA) in the early, mid, and late phases (D–F) reveals a parafoveal area of choroidal hyperfluorescence. Spectral-domain optical coherence tomography (SD-OCT) B-scans at the level of the leaking point (blue line) demonstrate subretinal fluid and a flat, irregular pigment epithelial detachment (FIPED) at baseline (G) and after several months of observation (H), with a magnified view highlighting a retinal pigment epithelium (RPE) break (red asterisk, I). OCT angiography (OCTA, J) and OCTA overlaid on FA (K) colocalize the choroidal neovascularization (CNV) with the leakage point (red arrow).

**Figure 2.**
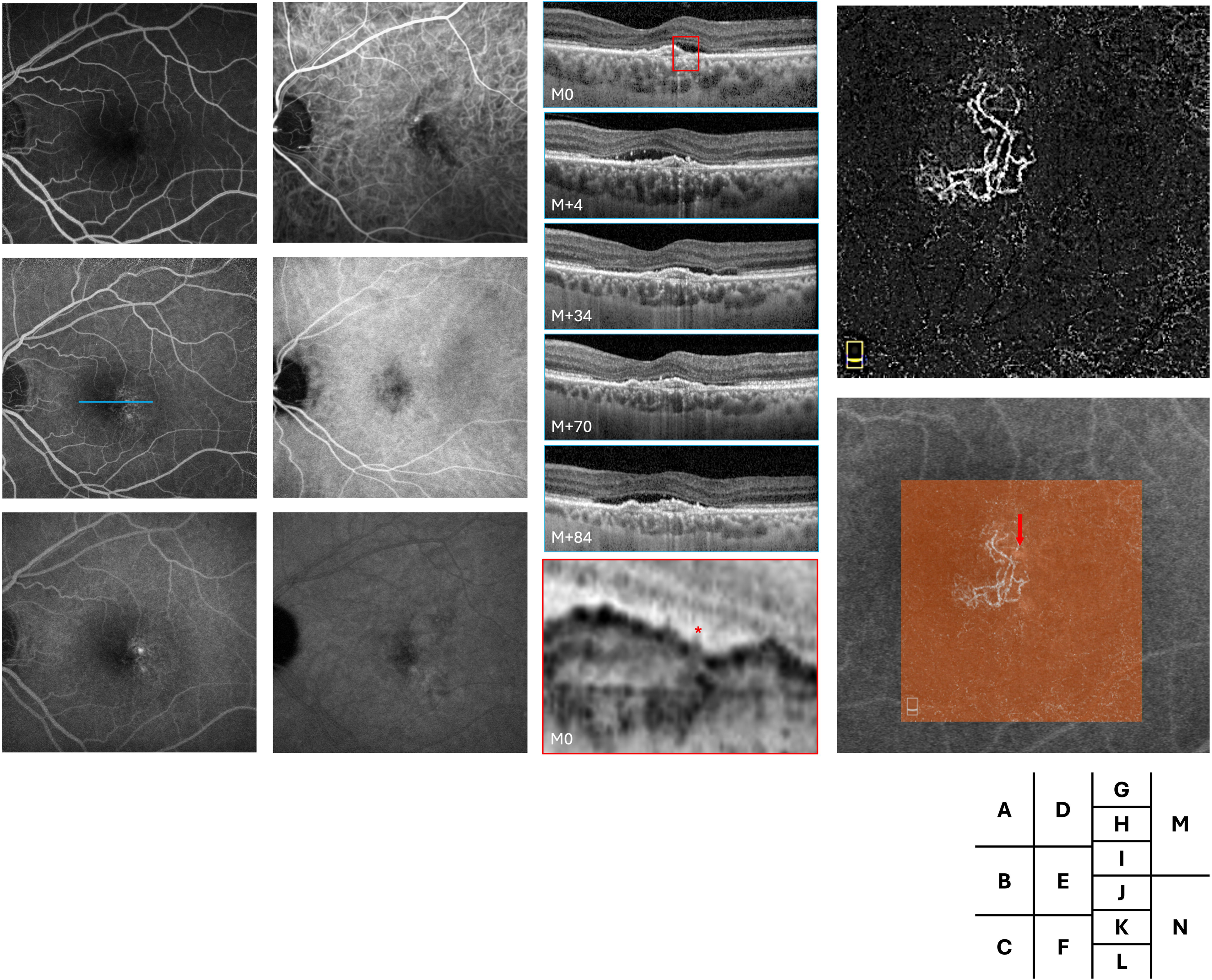
Multimodal follow-up imaging of the left eye in Case 2. Fluorescein angiography (FA) in the early, mid, and late phases (A–C) shows a retrofoveal focal leakage, while indocyanine green angiography (ICGA) in the early, mid, and late phases (D–F) reveals prominent pachyvessels. (G–L) Serial spectral-domain optical coherence tomography (SD-OCT) B-scans through the leakage site (blue line in B) illustrate disease evolution : 60 months before baseline (G, M–60), choroidal thickening with a flat, irregular pigment epithelial detachment (FIPED) and no subretinal fluid; 12 months before baseline (H, M–12), early FIPED formation; at baseline (I, M0, day of angiography), new subretinal fluid and photoreceptor outer segment elongation; 2 months later (J, M+2), increased fluid; 32 months later (K, M+32), persistent fluid despite three intravitreal anti-VEGF injections; and 66 months later (L, M+66), complete and durable resolution four months after initiating daily topical prednisone, maintained without recurrence for three years. OCT angiography (OCTA, M) and OCTA overlaid on FA (N) demonstrate the colocalized choroidal neovascularization (CNV) and leakage point (red arrow).

**Figure 3.**
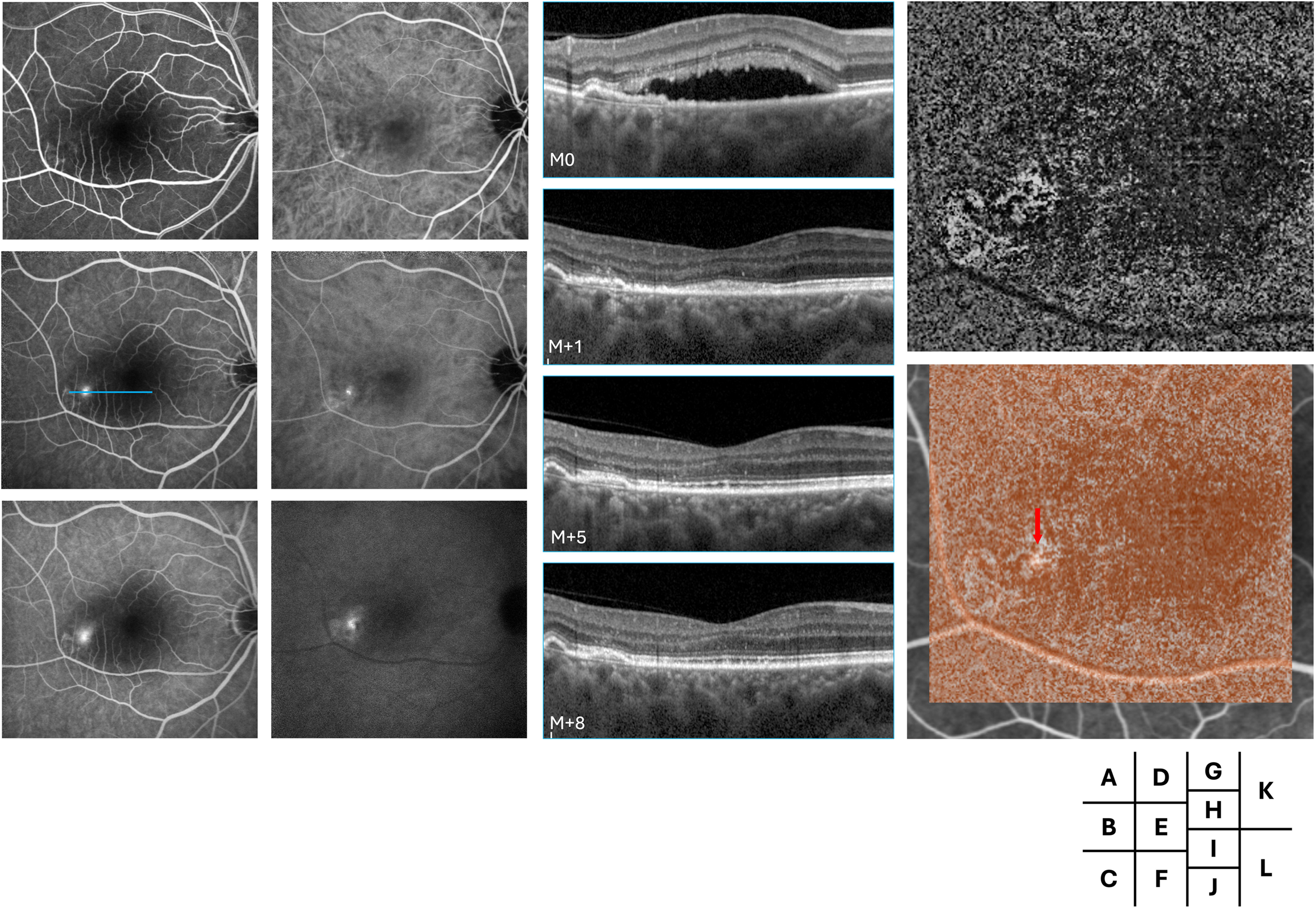
Multimodal follow-up imaging of the left eye in Case 3. Fluorescein angiography (FA) in the early, mid, and late phases (A–C) shows a focal hyperfluorescent leakage point, while indocyanine green angiography (ICGA) in the corresponding phases (D–F) reveals a parafoveal area of choroidal hyperfluorescence. Serial spectral-domain optical coherence tomography (SD-OCT) B-scans at the level of the leaking point (blue line, G–J) demonstrate subretinal fluid and a flat, irregular pigment epithelial detachment (FIPED) with associated fluid at presentation, which progressively resolves over time. Optical coherence tomography angiography (OCTA, K) and OCTA overlaid on FA (L) colocalize the choroidal neovascularization (CNV) with the leakage point (red arrow).

**Figure 4.**
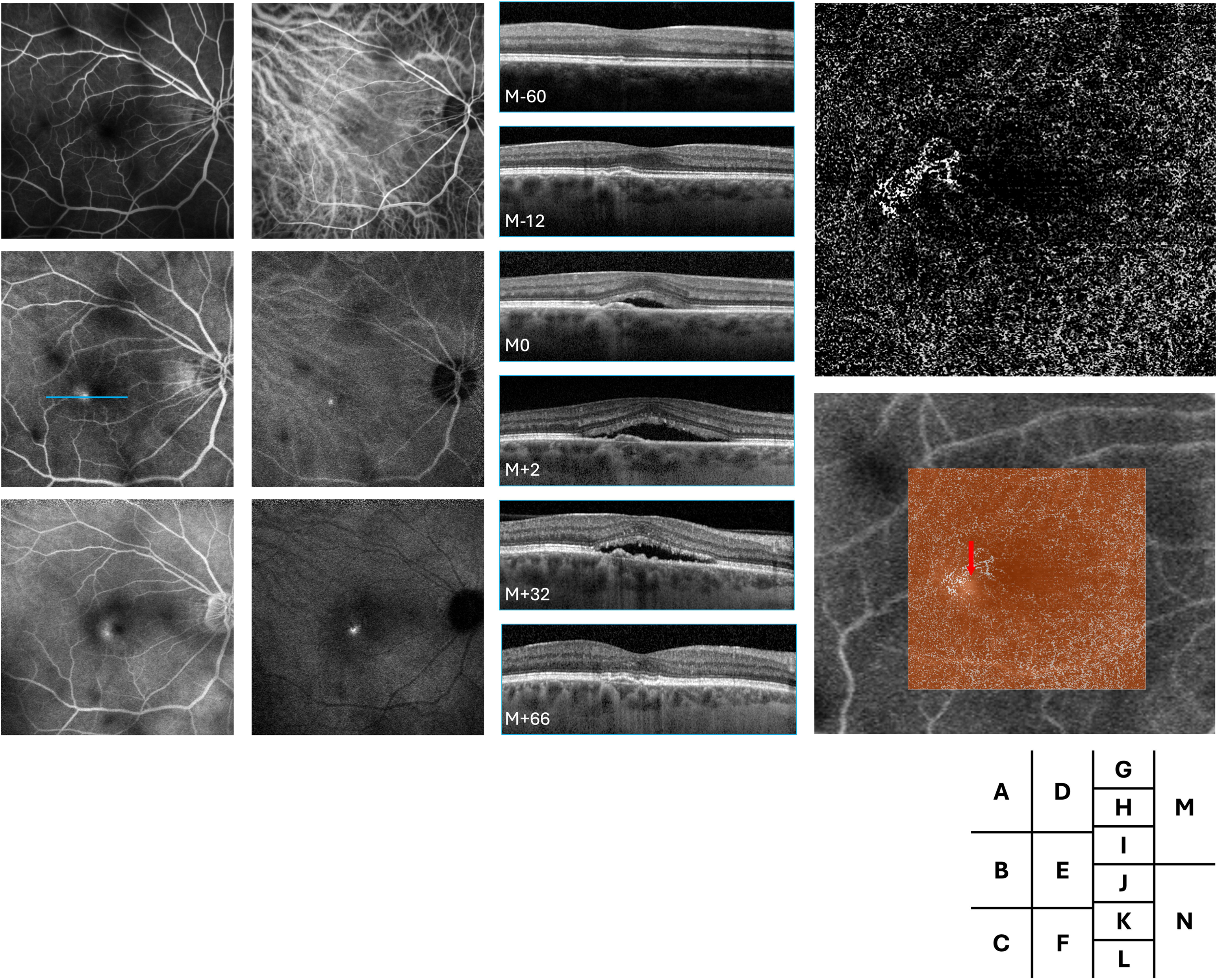
Multimodal follow-up imaging of the left eye in Case 4. Fluorescein angiography (FA) in the early, mid, and late phases (A–C) shows a subtle hyperfluorescent point, while indocyanine green angiography (ICGA) in the corresponding phases (D–F) reveals late hyperfluorescence at the leakage site. Spectral-domain optical coherence tomography (SD-OCT) B-scans at the level of the leaking point (blue line) demonstrate subretinal fluid and a flat, irregular pigment epithelial detachment (FIPED) at baseline (G) and after follow-up at 3 months, 2 years, 5 years, and 7 years (H–K), with a magnified view highlighting a retinal pigment epithelium (RPE) break (red asterisk, L). Optical coherence tomography angiography (OCTA, M) and OCTA overlaid on FA (N) colocalize the choroidal neovascularization (CNV) with the leakage point (red arrow).

### CNV and Leaking Points

In all cases, the CNV lesions were located close to the fovea, with three classified as retrofoveal (cases 1, 2, 4) and one as temporal (case 3). The morphology of the vessels varied: two patients had mature, “dead-tree” type vessels (Figures 1G, 4E) and two had immature “coral-like” or intermediate pattern (Figures 2E, 3E).

Each patient exhibited a single fluorescein angiography-confirmed leakage point. In all cases, the leakage site colocalized precisely with the neovascular lesion identified on OCTA. Multimodal imaging for each case, including FA and OCTA overlay, demonstrated the clear spatial association between the area of FA-confirmed leakage and the corresponding CNV lesion. Other signs of leakage were observed on SD-OCT such as lucency in cases 1 and 3, outer segment erosion above RPE in case 3 and RPE break in cases 1 and 4 (Figures 1I, 3, 4L).

### Timing of CSCR Onset and Serous Fluid Fluctuations

In three patients, CSCR was diagnosed at the same time as the CNV. In the remaining case, CSCR developed 12 months after the initial identification of CNV in a patient with FCE.

Fluctuations in subretinal fluid were observed in all four cases, occurring independently of anti-VEGF IVT injections. These changes were not temporally associated with injections, suggesting a mechanism not solely driven by VEGF-dependent CNV exudative activity. In one case (Case 4), the subretinal fluid persisted despite monthly anti-VEGF injections (Figure 4F–K).

## Discussion

This series of rare cases highlights a potential pathogenic link between CNV and the onset of acute CSCR in the context of pachychoroid. In these patients, FA revealed that the leaking point associated with and possibly responsible for subretinal fluid was located at the site of an RPE elevation above a CNV lesion. This spatial colocalization and the chronology of the events suggest that CNV may not merely be a late complication of chronic CSCR, but may instead act as a direct trigger for acute CSCR

CNV has been typically recognized as a complication of complex/ chronic CSCR(Hagag et al., 2020) in about 36% of cases and presenting as elevated RPE with double sign layer(Hagag et al., 2022). In a prospective longitudinal study, CNV complicated about 13% of CRSC, with higher risks for chronic cases, long-lasting fluid during the first year, older age and foveal involvement(Zhou et al., 2022). In the time course of chronic CSCR complicated by CNV, episodes of leakage can occur, often located in area of mid-phase ICG hyperpermeability, near or far from the neovessels and despite ongoing anti-VEGF treatment. It is therefore important to accurately locate the areas of leakage that could be treated by laser or focal PDT.(Feenstra et al., 2024) The multifactorial origin of the subretinal fluid in these cases could explain an incomplete response to anti-VEGF(Lejoyeux et al., 2022). On the other hand, Pang and Freund described ten years ago, the PNV phenotype as the presence of type 1 CNV underlying a shallow RPE elevation in eyes with thick choroid and no evidence of exudative detachment or autofluorescence changes suggestive of previous or actual CSCR.(Pang and Freund, 2015) The angiographic characteristics of PNV are a late fluorescein leakage with undetermined origin, and a typical late staining plaque on ICGA.(Pang and Freund, 2015) The clinical presentation of our cases does not correspond to any of these descriptions and appears rather as a simple and acute form of CRSC whose leakage point overlooks a neovessel. Although not formally described, this clinical form was suggested, as neovessels could be added signs in simple or complex forms of CRSC when we proposed reclassifying the disease.(Chhablani et al., 2020) The recognition of CNV-induced acute CSCR is important, as it may carry therapeutic implications and raise intriguing pathogenic hypotheses. From a diagnostic perspective, while SD-OCT is widely used in routine cases of acute or simple CSCR, it does not allow precise localization of leakage sites (as seen on FA) or direct visualization of CNV, which is better detected with OCT-A. However, the interpretation of OCT-A can be limited by signal attenuation in areas of subretinal fluid, making repeat imaging after fluid resolution often necessary. The best therapeutic option in this clinical presentation is uncertain and whether an initial combination of anti-VEGF and PDT would lead to more favourable outcome would need to be evaluated. PNV can be managed with either anti-VEGF or photodynamic therapy (PDT).(Tanaka et al., 2024) In cases where PNV is associated with CSCR-like features, treatment response may differ. Specifically, CSCR-predominant PNV has been associated with more favourable outcomes, including a lower recurrence rate of subretinal fluid following PDT or anti-VEGF therapy.(Maruyama-Inoue et al., 2024) In our cohort, several cases exhibited subretinal fluid fluctuations that appeared independent of anti-VEGF IVT. In such cases, targeted treatments—such as limited-spot PDT or possibly topical corticosteroids—may help reduce subretinal fluid and lower the risk of recurrence.(Kim et al., 2025; Zaman et al., 2025) However, the efficacy and safety of these approaches in this specific context remain to be established and warrant evaluation in prospective clinical studies.

The mechanism underlying the leakage point in these cases is particularly interesting. It closely resembles the leakage point observed above a large choroidal vessel adjacent to Bruch’s membrane, where the choriocapillaris is no longer identifiable. While mechanical factors may play a role—given that mechanical stress activates the YAP/TAZ pathway,(Totaro et al., 2018) which is crucial for maintaining RPE differentiation(Lu et al., 2020) —they cannot be considered solely responsible. Serous retinal detachment (SRF) without a detectable leakage point is frequently observed in situations involving mechanical stress, such as dome-shaped macula, large drusenoid PED, or choroidal tumors. In these instances, diffuse RPE dysfunction is the underlying cause of SRF.(Caillaux et al., 2013; Dansingani et al., 2016; Viggiano et al., 2025) We hypothesize that, in pachychoroid disease, vascular remodelling is associated with—and may even be secondary to—choroidal autonomic neuropathy.(Leclercq et al., 2023) Under physiological conditions, the entire vascular system is regulated by the autonomic nervous system (ANS), and RPE homeostasis and transport activity are modulated by neuropeptides and adrenergic signalling. Large nerves and arteries are typically distant from the RPE, thereby preventing uncontrolled catecholamine discharges from activating the RPE, despite the presence of adrenergic receptors on its surface. In pachychoroid disease, pachyvessels with pathological innervation—similar to the mature neovessels often observed in CSCR(Bousquet et al., 2018b)—may serve as a source of abnormal and uncontrolled neural activation due to their proximity to the RPE. From a pathophysiological perspective, dysfunctional vascular neural control may promote microthrombosis of the choriocapillaris, abnormal fibrin clotting, and RPE dysfunction, consistent with histological findings of RPE damage at leakage sites in monkeys.(Yoshioka et al., 1982; Yoshioka and Katsume, 1982b) Indeed, choroidal haemostasis is under adrenergic control(Gatica et al., 2023) and the sympathetic nervous system directly produces plasminogen activator around vessels.(O’Rourke et al., 2005) Ion transport across RPE in under adrenergic signalling, and excessive epinephrine can induce RPE cell apoptosis.(Sibayan et al., 2000) On the other hand, vasointestinal peptide (VIP) controls RPE homeostasis and barrier function.(Koh, 2000; Maugeri et al., 2017) Altogether, these mechanisms suggest that choroidal neuropathy, which may underlie the pachychoroid phenotype, combined with the close proximity of innervated vessels to the RPE, could induce RPE pump dysfunction, alter fibrinolysis, and activate matrix metalloproteinases (MMPs), thereby triggering the development of CSCR. This aligns with the view of CSCR as a maladaptive stress response. A schematic representation of this hypothesis is shown in **Figure 5**.

**Figure 5.**
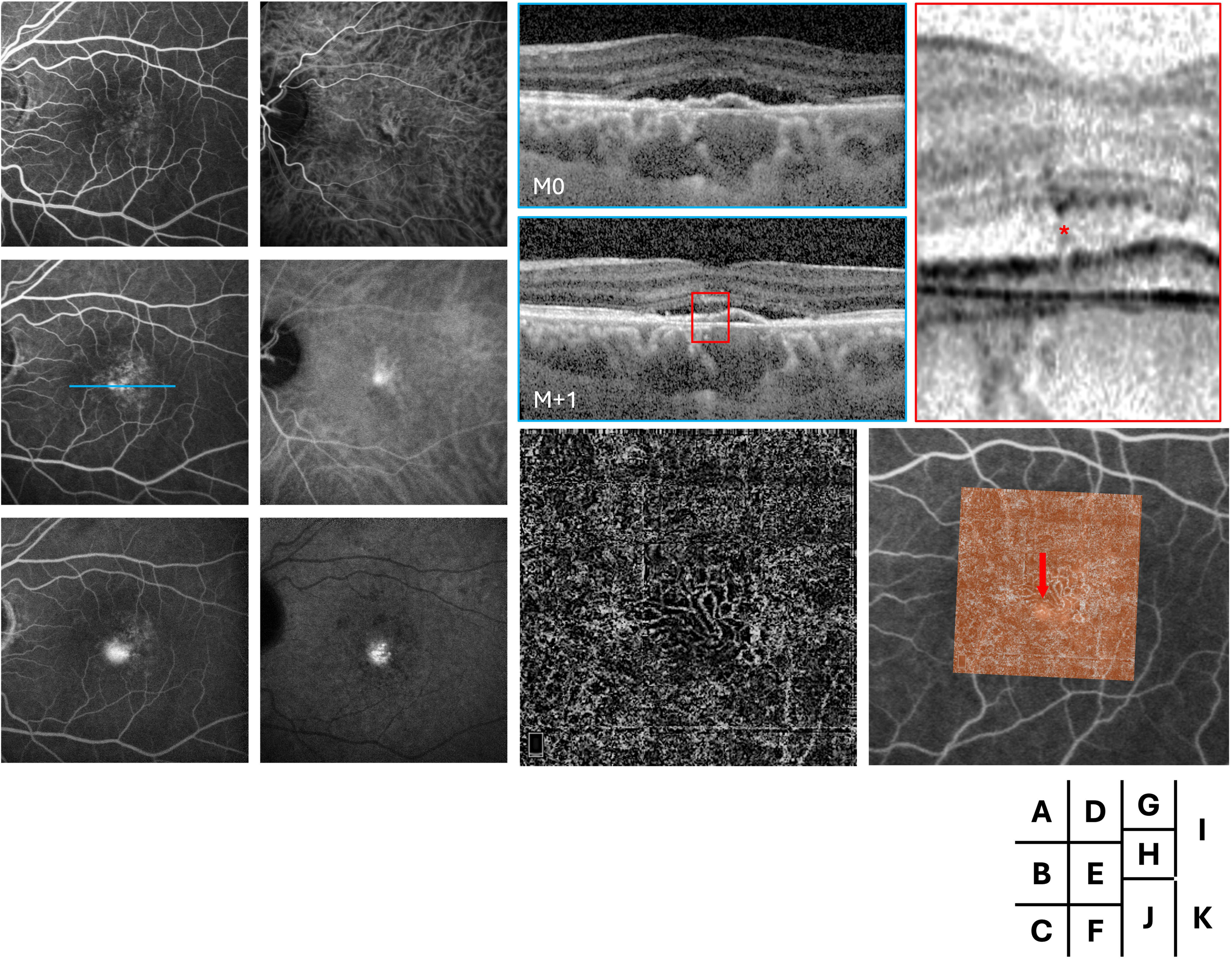
Schematic representation of the proposed hypothesis linking choroidal nerve signaling to leakage from choroidal neovascularization. Choroidal nerves release epinephrine, which activates matrix metalloproteinase-9 via VIP receptors on the retinal pigment epithelium (RPE). This disrupts RPE barrier integrity, facilitating fluid leakage. Simultaneously, choroidal microthrombosis contributes to local ischemia and inflammation, further promoting leakage.

This study has limitations. The retrospective design and small sample size limit statistical generalizability. OCTA may be subject to projection artifacts and false positives, particularly in cases with poor fixation or significant subretinal fluid. The chronology of event could be challenged as non-symptomatic episodes might have occurred without notice. Nevertheless, the consistent multimodal findings across cases support the concept that CNV may act as a triggering factor for CSCR under certain conditions.

Ultimately, these findings suggest that CSCR may occasionally result from a neurovascular dysfunction initiated by quiescent CNV or choriocapillaris injury—challenging the traditional view of CNV as a consequence rather than a cause of CSCR.

## Data Availability

All data produced in the present study are available upon reasonable request to the authors

---

**Supplemental Figure 1.** Multimodal imaging in acute and chronic central serous chorioretinopathy (CSCR).

Top row (A–D): Acute CSCR. (A) Spectral-domain optical coherence tomography (SD-OCT) shows serous retinal detachment with preservation of the outer retinal layers. (B) Fundus autofluorescence (FAF) reveals faint hyperautofluorescence in the macular region, corresponding to subretinal fluid. (C) Early-phase fluorescein angiography (FA) demonstrates focal “smokestack” leakage. (D) Indocyanine green angiography (ICGA) shows minimal retinal pigment epithelium (RPE) alteration.

Bottom row (E–H): Chronic CSCR. (E) SD-OCT reveals outer retinal atrophy, excavation and flat irregular pigment epithelial detachment. (F) FAF displays a mottled pattern of hypo- and hyperautofluorescence consistent with chronic RPE damage. (G) Early-phase fluorescein angiography (FA) shows multiple areas of diffuse leakage and window defects. (H) Late-phase ICGA (25 min) highlights extensive RPE atrophy and choroidal hyperpermeability.

**Supplemental Figure 2.** Multimodal imaging of the contralateral eyes. For each patient (from left to right: patients 1 to 4), the following modalities are shown: early-phase fluorescein angiography (FA, top left), early-phase indocyanine green angiography (ICGA, top right), late-phase FA (middle left), late-phase ICGA (middle right), blue autofluorescence (BAF, center), and foveal optical coherence tomography (OCT, bottom). In Patient 1, a quiescent choroidal neovascularization (CNV) is visible, with optical coherence tomography angiography (OCTA) superimposed on the BAF image to better localize the lesion.

