## Supplementary figures and images for "Choroidal Neovascularization as a Trigger for Central Serous Chorioretinopathy"

### Supplemental Figure 1

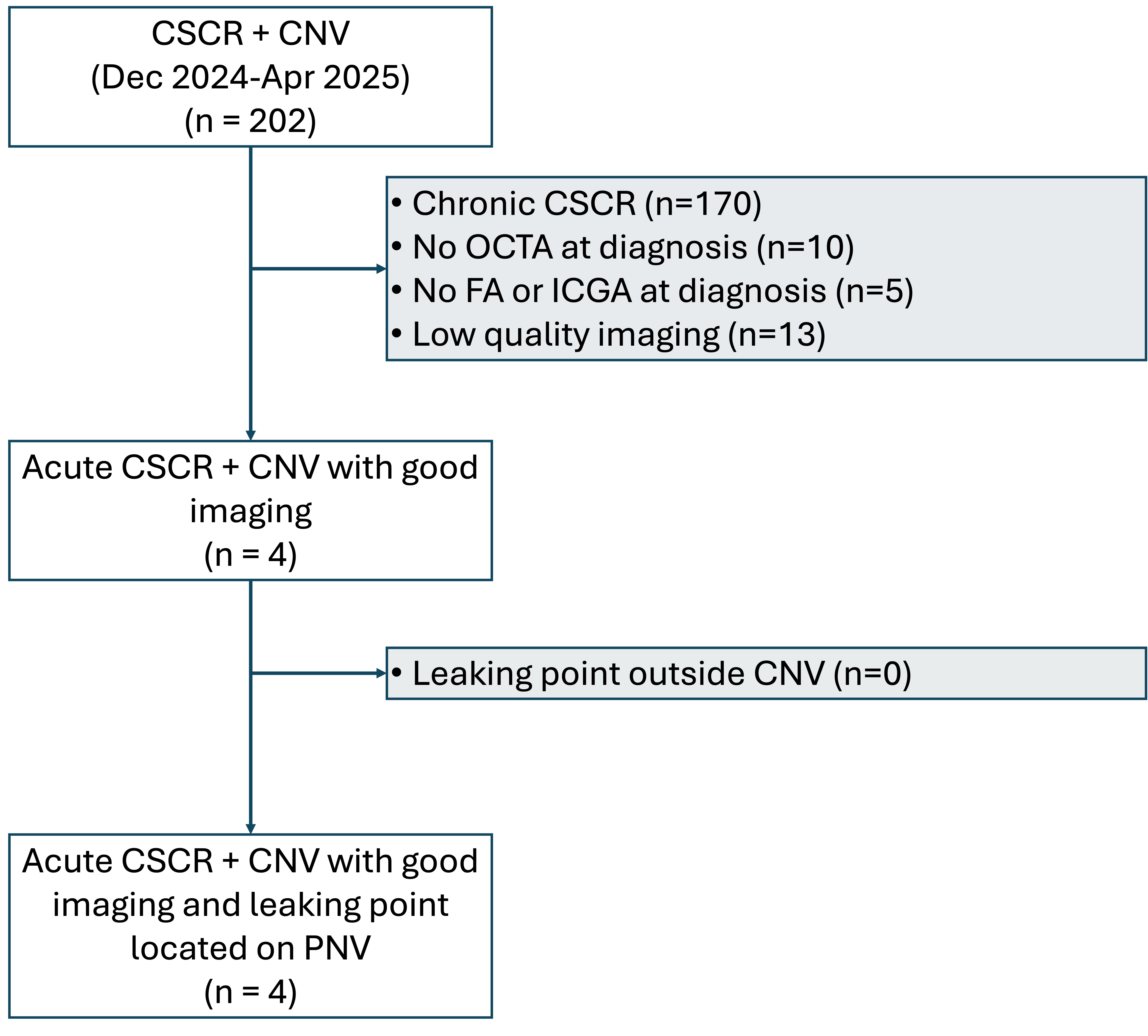

### Supplemental Figure 2

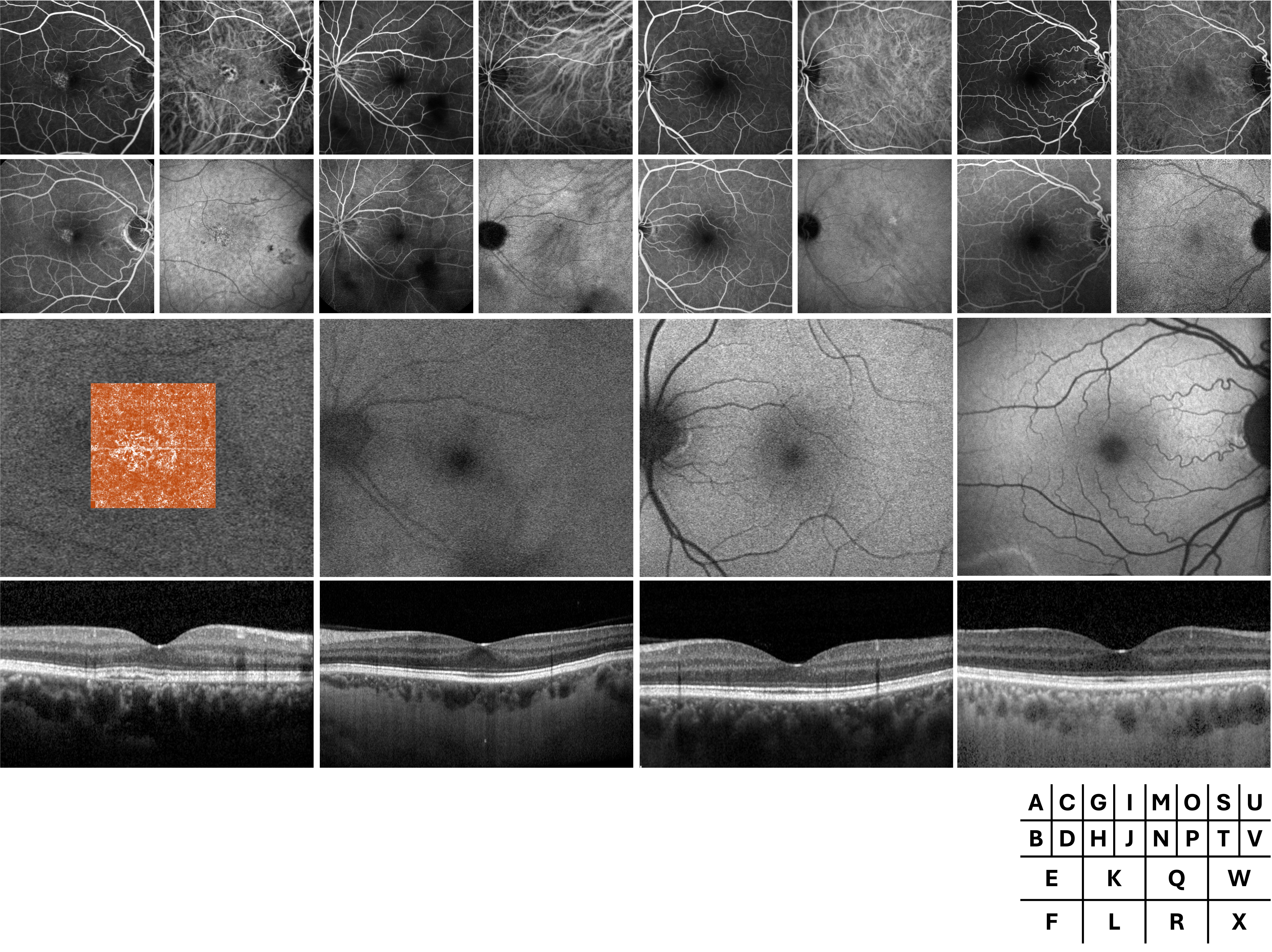
